# Hyperkalemia-related Heart Failure Therapy Discontinuation and the Association with Outcomes in Patients with Heart Failure

**DOI:** 10.1101/2025.03.06.25323518

**Authors:** Aanchel Gupta, Sunjidatul Islam, Douglas C. Dover, Padma Kaul, Finlay McAlister, Justin Ezekowitz

**Author notes:** Address for Correspondence: Justin Ezekowitz, MBBCh, MSc, 4-120 Katz Group Centre for Pharmacy and Health Research University of Alberta, Edmonton, AB, T6G 2E1.

## Abstract

**Background:** Renin-angiotensin-aldosterone system (RAAS) inhibitors are essential treatments for heart failure (HF) patients, but their use is often limited by hyperkalemia.

**Objective:** This study assesses the incidence of hyperkalemia in chronic HF patients on RAAS inhibitors, examines changes in therapy following hyperkalemia episodes, and evaluates the impact of RAAS inhibitor discontinuation or down-titration on patient outcomes.

**Methods:** We conducted a population-based cohort study of patients hospitalized or visiting the emergency department in Alberta for chronic HF from April 2012 to March 2020, focusing on those with RAAS inhibitor-associated hyperkalemia. Episodes of hyperkalemia (K >5.0 mmol/L) were monitored, and patients were followed for 30 days to determine if their RAAS therapy was maintained, reduced, or discontinued.

**Results:** Among 7527 HF patients, we identified 123,038 RAAS inhibitor treatment years, resulting in 17 hyperkalemia events per 100 treatment years. Hyperkalemia led to RAAS inhibitor discontinuation in 35.2% of cases, down-titration in 8.4%, and continuation in 56.4%. Discontinuation or down-titration was more common when serum potassium exceeded 6.0 mmol/L (49.4%) compared to lower levels. Over a median follow-up of 1.4 years, discontinuing or down-titrating RAAS inhibitors was associated with increased all-cause mortality (aHR 1.80), higher cardiovascular hospitalizations (aHR 1.09), and more frequent ED visits for HF (aHR 1.17) compared to continued therapy.

**Conclusions:** Discontinuation or down-titration of RAAS inhibitors in HF patients is associated with higher mortality and cardiovascular events. Strategies to maintain RAAS therapy after hyperkalemia episodes may improve patient outcomes.

## Introduction

Heart failure (HF) is a major public health problem, with frequent emergency department (ED) visits, hospital admissions, and a 5-year survival rate of roughly 50%.^1^ The use of guideline-directed medical therapy (GDMT) improves clinical outcomes in patients with chronic HF.^1–3^ Nevertheless, electrolyte imbalances such as hyperkalemia are frequently observed in patients receiving HF therapies such as renin-angiotensin-aldosterone system inhibitors (RAASi) including angiotensin-converting enzyme inhibitors (ACEi),^4–9^ angiotensin receptor blockers (ARB),^10–13^ angiotensin receptor-neprilysin inhibitors (ARNI), and mineralocorticoid receptor antagonists (MRA).

Hyperkalemia, defined as serum potassium levels greater than 5 mmol/L (or 5 mEq/L), occurs in up to 40% of patients with chronic HF, and is an independent predictor of arrhythmias, hospitalizations, and mortality.^14,15^ The European Society of Cardiology (ESC) issued an expert consensus statement on the optimal management of patients with cardiovascular disease and incident hyperkalemia in 2018 and recommended temporary discontinuation of RAASi with serum potassium levels above 6 mmol/L, and watchful continuation of RAASi in levels between 5 and 6 mmol/L.^14^ They recommended that if a short-term cessation of RAASi is deemed necessary, RAASi should be carefully reintroduced as soon as possible while monitoring potassium levels.^16^ Prior estimates of hyperkalemia following MRA initiation may not be informative as most were based solely on administrative data rather than laboratory data.

Despite the availability of other management options, clinicians often respond to hyperkalemia with discontinuation or down-titration of beneficial treatments such as ACEi/ARBs/ARNIs or MRAs.^17–22^ In the ESC Heart Failure Long-Term Registry of 12,440 patients with HF with reduced ejection fraction (HFrEF), hyperkalemia was the reason for the non-use of ACEi/ARB and MRA in 8.5% ^23^ and 35.1% of patients, respectively.^24^ Despite their established mortality benefits, unfortunately, these medications are rarely reintroduced after discontinuation.^18,19,25–29^

We conducted a population-based cohort study in a single geographic region of 4.7 million people to investigate the incidence of hyperkalemia among patients with chronic HF receiving RAASi treatment, the change of HF therapies following hyperkalemia, and the association between RAASi discontinuation or down-titration and patient outcomes in patients presenting with HF and hyperkalemia.

## Methods

### Data Sources

Anonymized data for the study was retrieved from the comprehensive administrative databases maintained by Alberta Health Services and Alberta Health.^30,31^ These databases contained data on all patient interactions with the health care system anywhere in the province. The databases are comprehensive and are linked by a unique lifetime identifier (ULI), unique to each patient in Alberta. The databases were merged into a relational database to assemble a cohort of adult patients with HF who resided in Alberta between April 01, 2002 and March 31, 2020. The resulting integrated database allowed us to track patients longitudinally from their index HF encounters during the follow-up period.

National Ambulatory Care Reporting System (NACRS) contains records of every visit a patient makes to an ED or ambulatory care center situated within an acute care facility. Data from NACRS was merged with inpatient data from the hospital discharge database (Discharge Abstract Database [DAD]) to identify admissions to the hospital, in-hospital outcomes, procedures and other codes related to comorbid conditions, derived from previous hospitalizations. The information in these databases has been demonstrated to be highly accurate for use in the research setting^30–32^ and codes diagnoses using the International Classification of Disease, 10th revision (ICD-10) and procedures with the Canadian Classification of Health Intervention (CCI) codes. Primary care visits were captured through the Physicians Claims database, which uses ICD-9 codes for diagnoses. Data on medications were captured using Pharmaceutical Information Network (PIN) data, which identifies prescription drug information (Drug Identification Number (DIN), Anatomical Therapeutic Chemical Classification (ATC), dispensing date, amount, and the number of days of supply) for all medications dispensed in Alberta from April 1, 2008 onwards, regardless of patient age.

Laboratory studies were captured through a province-wide laboratory repository that has captured all inpatient, ED and outpatient laboratory results since April 1, 2012. ^33,34^ Echocardiograms are from the two tertiary care hospitals in the province.^35^

### Ethics and Data Sharing

The study was approved by the Health Research Ethics Board of the University of Alberta. The data that support the findings of this study are available from the Alberta Health Services, but restrictions apply to the availability of these data, which were used under a data-disclosure agreement for the current study and are not publicly available. To request authorization to obtain these data by direct access, contact.

### Study Design and Population Selection

For this retrospective cohort study, Albertan adults who were hospitalized or had ED visits with HF as the primary diagnosis (ICD-10 code I50.x) between April 1, 2002 and March 31, 2020 and were alive on April 1, 2008, were identified. Of these, the study population consisted of patients who had at least one dispensation for any RAASi (ACEi, ARBs, ARNIs, or MRAs) and developed hyperkalemia between April 1, 2012 and March 31, 2020.

### Identification of RAASi-induced Hyperkalemia

All hyperkalemia events (defined as serum potassium >5.0 mmol/L) experienced by HF patients between the April 2012 and March 2020 fiscal years were identified. In the case of multiple measurements on the same day, the highest potassium value was used. The severity of hyperkalemia was stratified into three categories: mild (>5.0 to <5.5 mEq/L), moderate (5.5 to 6.0 mEq/L), and severe (>6.0 mEq/L)^14^. All RAASi dispenses in the 30 days before the occurrence of a hyperkalemia event were identified. The most recent dispense was selected as the potential cause of hyperkalemia. We excluded hyperkalemia events with no associated RAASi dispenses. A hyperkalemia episode begins with the first hyperkalemia event; the episode ends at the first RAASi dispense within 30 days if there is one, otherwise the later of the first hyperkalemia event or the latest of other hyperkalemia events within 30 days of the first event. The value of the first hyperkalemia event was chosen as the potassium value for the episode. The first hyperkalemia episode of a patient related to RAASi was considered the index episode.

### Exclusion Criteria

To identify chronic HF patients, patients who had a HF hospitalization or ED visit in the six months before the index hyperkalemia episode were excluded. Additionally, patients who had a history of end-stage renal disease, kidney transplant, or hemodialysis in the past five years and who had potassium level >10 mmol/L (to avoid spurious values) were excluded from the analysis. With regards to excluding patients with end-stage renal disease, although this definition can vary, patients may be more likely to discontinue RAASi due to changes in kidney function during our follow-up period; reasons for discontinuation are not available in the data set and therefore these patients were preemptively excluded.

### Dose Intensity Calculation for RAASi

Treatment changes were evaluated from the last day of the episode (possibly defined by RAASi dispenses) to 30 days later. All RAASi dispenses on the first day with any RAASi dispenses were used to categorize treatment as same-dose, increased dose, or down titration. The dose intensity score was computed to explore the changes in the dosage of RAASi between the dispense responsible for the occurrence of hyperkalemia and the subsequent dispense aimed at managing the hyperkalemia event. The total daily dosage (TDD) of each medication was calculated by dividing the dispensation amount (number of tablets) by the number of days of supply and then multiplying it by the strength of the medication. The TDD of ACEis, ARBs, and MRAs were converted to the lisinopril, losartan, and spironolactone equivalent dosage, respectively. The equivalent dosages were assigned a dose intensity score according to Supplementary Table 1. The dose intensity score ranged from 0 to 5 for ACEis, ARBs, and ARNIs and from 0 to 4 for MRAs. In the case of a combination or multiple medications from different drug classes dispensed on the same day, the dose intensity score for each medication was summed up to get the final score.

Patients without a RAASi dispense were classified as continuing on the same dose if the duration of the pre-episode dispense overlapped with the treatment period. Otherwise, patients without a RAASi dispense were classified as discontinuing.

### Other Variables

Patient characteristics were assessed at each RAASi-induced hyperkalemia episode, including age, sex, diabetes, hypertension, dyslipidemia, stroke, renal disease, coronary artery disease, and atrial fibrillation. Patient comorbidities were identified using ICD codes (Supplementary Table 2)^36,37^ from any hospitalizations, ambulatory encounters, or from two physician’s claims in the outpatient setting (at least 30 days apart within one year) in up to five years before the index episode. Using the provincial echocardiography databases, HF subtypes were identified including HFrEF (<40%), HF with midrange ejection fraction (HFmEF) (40-49%), and HF with preserved ejection fraction (HFpEF) (≥50%). In the case of multiple echocardiography studies, the left ventricular ejection fraction (LVEF) value from the echocardiography assessment closest to the index episode date was used. The most recent tests of sodium, serum creatinine, estimated glomerular filtration rate (eGFR), and hemoglobin were obtained if they were done between 7 days before and 2 days following the start of hyperkalemia episode. Medications used in the year before the hyperkalemia episode were extracted from the PIN database using the ATC codes (Supplementary Table 2).

### Outcomes

The clinical outcomes of mortality (all-cause), hospitalization (all-cause, cardiovascular-, or HF-related), and ED visits (all-cause, cardiovascular-, or HF-related), including the time to first event, were evaluated during the follow-up period. Cardiovascular disease and HF have been defined using the ICD-10 code “I” and “I50.x” as the major diagnoses, respectively. The secondary outcome was the occurrence of recurrent hyperkalemia defined as a potassium value >5.0 mmol/L. Patients were followed for the occurrence of outcomes from the latter of the episode end date or RAASi treatment until they had another RAASi-induced hyperkalemia event. In all outcomes, patients were followed until they moved out of province, died, or the study ended, whichever occurred first within two years.

### Statistical Analysis

Categorical variables were summarized as frequencies with proportions and compared across groups using chi-square tests. Continuous variables were presented as median with interquartile range (IQR) and compared with Kruskal Wallis tests. Factors associated with RAASi discontinuation or down-titration were identified using the binary logistic regression with the Generalized Estimating Equation (GEE) model. The multivariable model included age (linear spline with two knots at 65 and 75 years), sex, urban/rural residence, hospital type, HF type, the severity of hyperkalemia, dose intensity at hyperkalemia event (linear spline with one knot at 5), diabetes, hypertension, hyperlipidemia, coronary artery disease, atrial fibrillation, chronic renal disease, acute kidney injury, asthma, chronic obstructive pulmonary disease (COPD), anemia, cancer, depression and dementia.

The crude rate per 100 person-years (PY) was used to summarize the outcomes-all-cause mortality, hospitalizations (for any cause, cardiovascular, and HF), ED visits (for any cause, cardiovascular, and HF) and recurrent hyperkalemia. The rate was calculated by dividing the total number of outcomes that happened while patients remained in a particular treatment status by the duration of that treatment status throughout the follow-up period. To assess the outcome, patients were followed from the management of RAASi-associated hyperkalemia episode to the next hyperkalemia episode, death, migration out of province or the end of the study, whichever occurred first within two years. We performed the time-varying Cox regression analysis using the inverse probability weight (IPW) to estimate the hazard ratio for RAASi discontinuation or down-titration compared with continuation. Inverse probability was calculated as the average treatment effect in the treated (ATT) using the covariate balancing propensity score approach, which ensures that variables are balanced between treatment groups.^38^ The model included age, sex, residence, hospital type, HF type, the severity of hyperkalemia, dose intensity associated with hyperkalemia event, diabetes, hypertension, hyperlipidemia, coronary artery disease, atrial fibrillation, chronic renal disease, acute kidney injury, asthma, COPD, anemia, cancer, depression and dementia. For non-death outcomes, death was considered as a competing risk and a cause-specific Cox regression model was used. A two-sided p-value <0.05 was considered statistically significant. All analyses were performed using the SAS statistical software version 9.4 (SAS Institute Inc.).

## Results

### Baseline Characteristics

The study identified 67,557 adults who were hospitalized or visited ED with HF between April 1, 2002 and March 31, 2020. As of April 1, 2012, a total of 54,025 patients were alive, with about 75% of them having a dispensation for any RAASi between April 1, 2012, and March 31, 2020. Out of the patients who received a RAASi dispensation, a total of 17,531 (43.3%) patients experienced RAASi-associated hyperkalemia (episodes n=54,419) (Supplementary Figure 1). Of these, patients (n=8240) who had an HF hospitalization or ED visit in 6 months before their first hyperkalemic episode and patients (n=1764) who had a history of end-stage renal disease or kidney transplant or dialysis were not included in the analysis. The study cohort finally consisted of 7527 adults with chronic HF who experienced a total of 21,026 episodes of hyperkalemia associated with RAASi. This contributed to a rate of 17 hyperkalemia episodes per 100 RAASi-treatment-years (over 123,038 RAASi-treatment-years) with follow-up starting from the first RAASi dispensation during the study period. Table 1 presents the characteristics of HF patients who experienced RAASi-associated hyperkalemia events.

**Table 1.** Characteristics of patients with heart failure at hyperkalemia episodes according to their treatment status.

| Variable Label | Statistic | Discontinuation | Down-titration | Up-titration | Same Dose | Total | p-value |
| --- | --- | --- | --- | --- | --- | --- | --- |
| Total N | N (%) | 7394 (35.2) | 1766 (8.4) | 1219 (5.8) | 10,647 (50.6) | 21,026 (100.0) |  |
| Age | Median (IQR) | 78 (69, 85) | 78 (70, 86) | 76 (68, 84) | 80 (70, 87) | 79 (70, 86) | <0.0001 |
| Sex |  |  |  |  |  |  |  |
| Females | n (%) | 3028 (41.0) | 801 (45.4) | 491 (40.3) | 5210 (48.9) | 9530 (45.3) | <0.0001 |
| Males | n (%) | 4366 (59.0) | 965 (54.6) | 728 (59.7) | 5437 (51.1) | 11,496 (54.7) |  |
| Residence |  |  |  |  |  |  |  |
| Rural | n (%) | 1687 (22.8) | 394 (22.3) | 314 (25.8) | 2522 (23.7) | 4917 (23.4) | 0.0767 |
| Urban | n (%) | 5707 (77.2) | 1372 (77.7) | 905 (74.2) | 8125 (76.3) | 16,109 (76.6) |  |
| HF Type |  |  |  |  |  |  | <0.0001 |
| rEF | n (%) | 1223 (16.5) | 367 (20.8) | 301 (24.7) | 1420 (13.3) | 3311 (15.7) |  |
| mEF | n (%) | 443 (6.0) | 107 (6.1) | 63 (5.2) | 647 (6.1) | 1260 (6.0) |  |
| pEF | n (%) | 1528 (20.7) | 353 (20.0) | 185 (15.2) | 2307 (21.7) | 4373 (20.8) |  |
| Missing | n (%) | 4200 (56.8) | 939 (53.2) | 670 (55.0) | 6273 (58.9) | 12,082 (57.5) |  |
| Diabetes | n (%) | 1719 (23.2) | 436 (24.7) | 301 (24.7) | 2989 (28.1) | 5445 (25.9) | <0.0001 |
| Hypertension | n (%) | 2076 (28.1) | 523 (29.6) | 348 (28.5) | 3244 (30.5) | 6191 (29.4) | 0.0058 |
| Hyperlipidemia | n (%) | 237 (3.2) | 43 (2.4) | 30 (2.5) | 328 (3.1) | 638 (3.0) | 0.2279 |
| CAD | n (%) | 1537 (20.8) | 414 (23.4) | 265 (21.7) | 2261 (21.2) | 4477 (21.3) | 0.1039 |
| PVD | n (%) | 187 (2.5) | 46 (2.6) | 33 (2.7) | 289 (2.7) | 555 (2.6) | 0.8934 |
| Atrial fibrillation | n (%) | 832 (11.3) | 228 (12.9) | 146 (12.0) | 1194 (11.2) | 2400 (11.4) | 0.1790 |
| Stroke/TIA | n (%) | 67 (0.9) | 22 (1.2) | 15 (1.2) | 150 (1.4) | 254 (1.2) | 0.0259 |
| Liver disease | n (%) | 108 (1.5) | 26 (1.5) | 16 (1.3) | 178 (1.7) | 328 (1.6) | 0.5898 |
| Chronic renal disease | n (%) | 574 (7.8) | 139 (7.9) | 84 (6.9) | 995 (9.3) | 1792 (8.5) | 0.0002 |
| Asthma | n (%) | 252 (3.4) | 63 (3.6) | 39 (3.2) | 320 (3.0) | 674 (3.2) | 0.3770 |
| COPD | n (%) | 929 (12.6) | 215 (12.2) | 150 (12.3) | 1618 (15.2) | 2912 (13.8) | <0.0001 |
| Anemia | n (%) | 281 (3.8) | 77 (4.4) | 52 (4.3) | 644 (6.0) | 1054 (5.0) | <0.0001 |
| Cancer | n (%) | 208 (2.8) | 57 (3.2) | 42 (3.4) | 265 (2.5) | 572 (2.7) | 0.0884 |
| Depression | n (%) | 367 (5.0) | 124 (7.0) | 65 (5.3) | 834 (7.8) | 1390 (6.6) | <0.0001 |
| Dementia | n (%) | 149 (2.0) | 47 (2.7) | 26 (2.1) | 278 (2.6) | 500 (2.4) | 0.0548 |
| Smoking | n (%) | 268 (3.6) | 53 (3.0) | 59 (4.8) | 420 (3.9) | 800 (3.8) | 0.0479 |
| Charlson Comorbidity Index | Median (IQR) | 5 (3, 7) | 5 (3, 7) | 5 (3, 7) | 5 (4, 7) | 5 (3, 7) | <0.0001 |
| **Laboratory Values** |  |  |  |  |  |  |  |
| Potassium value | Median (IQR) | 5 (5, 6) | 5 (5, 6) | 5 (5, 5) | 5 (5, 5) | 5 (5, 6) | <0.0001 |
| Severity of hyperkalemia |  |  |  |  |  |  |  |
| >5.0-<5.5 | n (%) | 5216 (70.5) | 1207 (68.3) | 938 (76.9) | 8068 (75.8) | 15,429 (73.4) | <0.0001 |
| 5.5-6.0 | n (%) | 1789 (24.2) | 491 (27.8) | 252 (20.7) | 2278 (21.4) | 4810 (22.9) |  |
| >6.0 | n (%) | 389 (5.3) | 68 (3.9) | 29 (2.4) | 301 (2.8) | 787 (3.7) |  |
| Serum creatinine value | Median (IQR) | 125 (97, 166) | 127 (100, 162) | 120 (96, 153) | 126 (98, 162) | 125 (98, 163) | 0.0072 |
| eGFR value | Median (IQR) | 43 (31, 60) | 43 (31, 58) | 46 (34, 60) | 42 (31, 57) | 43 (31, 59) | <0.0001 |
| Sodium value | Median (IQR) | 139 (136, 141) | 139 (135, 141) | 139 (136, 141) | 139 (136, 141) | 139 (136, 141) | 0.0323 |
| Hemoglobin, g/dL | Median (IQR) | 12 (11, 14) | 12 (11, 14) | 12 (11, 14) | 12 (11, 14) | 12 (11, 14) | 0.0140 |
| **Medications** |  |  |  |  |  |  |  |
| ACEi | n (%) | 5021 (67.9) | 1226 (69.4) | 868 (71.2) | 7330 (68.8) | 14,445 (68.7) | 0.0997 |
| ARB | n (%) | 2480 (33.5) | 628 (35.6) | 457 (37.5) | 3454 (32.4) | 7019 (33.4) | 0.0006 |
| MRA | n (%) | 3457 (46.8) | 839 (68.8) | 1214 (68.7) | 4849 (45.5) | 10,359 (49.3) | <0.0001 |

HF, heart failure; rEF, reduced ejection fraction; mEF, midrange ejection fraction; pEF, with preserved ejection fraction; CAD, coronary artery disease; PVD, peripheral vascular disease; TIA, transient ischemic attack; COPD, chronic obstructive pulmonary disease; eGFR, estimated glomerular filtration rate; ACEi, angiotensin-converting-enzyme inhibitors; ARB, angiotensin 2 receptor blockers; MRA, mineralocorticoid receptor antagonists

**Table 2.** Crude rates of outcomes at two years according to treatments.

|  | **Continuation**<br>**(n=10,647 patients)** |  | **Discontinuation/down-titration**<br>**(n=9160 patients)** |  |
| --- | --- | --- | --- | --- |
|  | **n** | **rate/100-PY** | **n** | **rate/100-PY** |
| **All-cause mortality** | 830 | 18.8 (17.6-20.2) | 1697 | 32.2 (30.7-33.8) |
| **Hospitalizations** |  |  |  |  |
| **Any cause** | 5308 | 123.7 (120.5-127.1) | 6147 | 126.2 (123.1-129.4) |
| **Cardiovascular diseases** | 1292 | 29.6 (28.0-31.3) | 1733 | 34.7 (33.1-36.4) |
| **Heart failure** | 807 | 18.4 (17.2-19.7) | 1059 | 21.0 (19.8-22.3) |
| **ED visits** |  |  |  |  |
| **Any cause** | 11,605 | 270.2 (265.3-275.1) | 12,228 | 252.5 (248.1-257.1) |
| **Cardiovascular diseases** | 1638 | 37.7 (35.9-39.6) | 2119 | 42.6 (40.9-44.5) |
| **Heart failure** | 922 | 21.1 (19.8-22.5) | 1320 | 26.3 (24.9-27.7) |
| **Recurrent hyperkalemia** | 8632 | 186.6 (182.7-190.6) | 9338 | 199.6 (195.6-203.7) |

PY, patient-years; ED, emergency department

Patients experiencing RAASi-associated hyperkalemia had a median age of 79 years (IQR 70, 86), and were more likely to be male (54.7%) and from urban locale (76.6%). Of all RAASi-associated hyperkalemia episodes, 73.4% had serum potassium <5.5 mEq/L, 22.9% had serum 5.5 to 6.0 mEq/L, and 3.7% had potassium >6.0 mEq/L. Among 42.5% of all patients with documented LVEF measurements, 37.0% had HFrEF, 14.1% had HFmEF, and 48.9% had HFpEF.

### Management of RAASi-associated Hyperkalemia

The development of hyperkalemia led to the discontinuation of RAASi in 35.2% of episodes, down-titration in 8.4%, and continuation in 56.4% (Figure 1). Discontinuation and down-titration of RAASi were more common than continuation when serum potassium exceeded 6.0 mEq/L (5.3% and 3.9% vs 2.8%) and when serum potassium was 5.5-6 mEq/L (24.2% and 27.8% vs 21.3%) (Table 1). Figure 2 shows the adjusted odds ratio (aOR) and 95% confidence interval (CI) for factors associated with discontinuation or down-titration of RAASi. Following adjustment, discontinuation or down-titration was less likely among patients aged above 75 years (per 5-year increase: aOR 0.89 95% CI 0.85-0.92, p<0.001). After a hyperkalemia episode, males (aOR 1.20, 95% CI 1.09-1.32, p<0.001), patients with moderate or severe hyperkalemia (aOR (1.37, 95%CI 1.28-1.47, p<0.001), and patients with asthma (aOR 1.30, 95% CI 1.04-1.62, p=0.021) were more likely to have their RAASi discontinued or down-titrated. Patients with diabetes, anemia, COPD and depression were less likely to have their RAASi discontinued or down-titrated after an episode of hyperkalemia.

**Figure 1.**
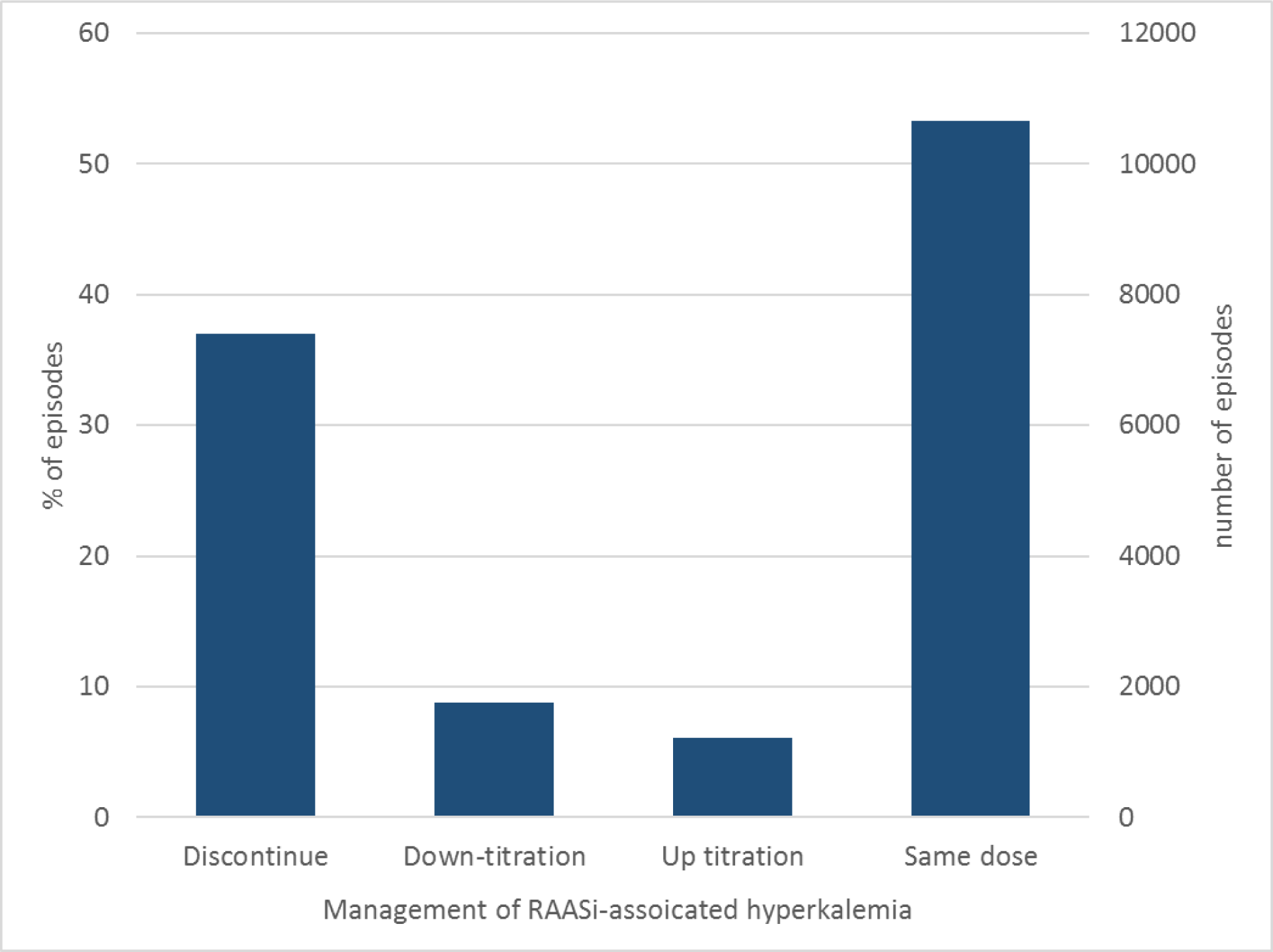
Management of RAASi-associated hyperkalemia Bar chart of the proportion (%) and number of episodes of hyperkalemia and changes in dosing of RAASi within 30 days of the hyperkalemia episode. RAASi, renin-angiotensin-aldosterone system inhibitors

**Figure 2.**
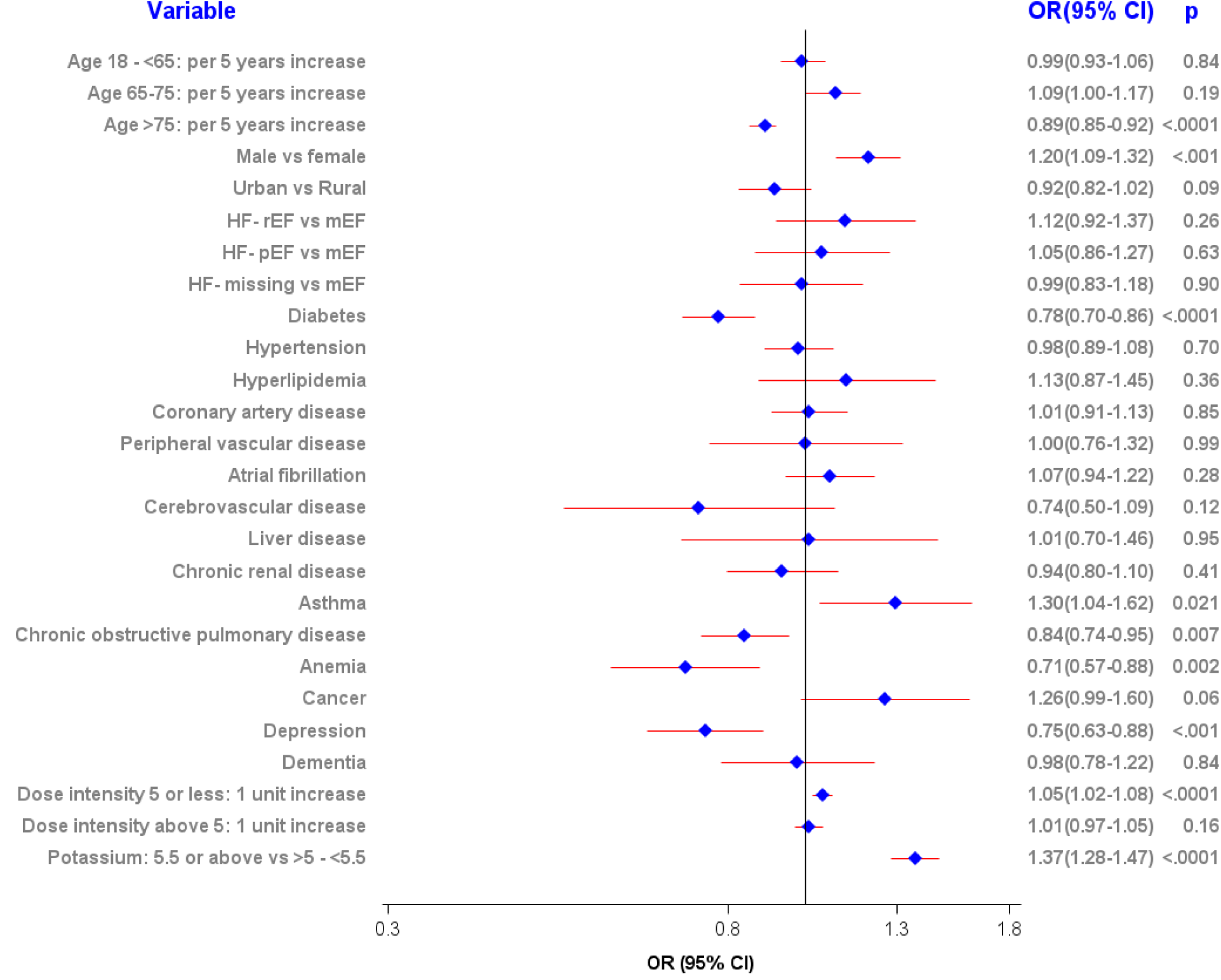
Factors associated with discontinuation or down-titration of RAASi after hyperkalemia Forest plot of adjusted ORs for factors associated with discontinuation or down-titration of RAASi within 30 days of the hyperkalemia episode. RAASi, renin-angiotensin-aldosterone system inhibitors; HFpEF, heart failure with preserved ejection fraction; HFrEF, heart failure with reduced ejection fraction; OR, odds ratio; CI, confidence interval

### RAASi Management and Outcomes

Table 3 presents the crude rate of outcomes (rates per 100 PY) and Figure 3 presents the weighted hazard ratios (aHR) and 95% CI of clinical outcomes computed from the time-varying Cox regression model employing inverse probability weighting. Over a median follow-up of 1.4 (IQR 0.7-2.0) years, discontinuation or down-titration of RAASi was associated with the increased risk of all-cause mortality (aHR 1.80, 95% CI 1.65-1.95, p<0.001), hospitalizations for cardiovascular disease (aHR 1.09, 95% CI 1.01-1.18, p=0.022), and ED visits for HF (aHR 1.17, 95% CI 1.07-1.27, p<0.001) as compared to the continuation of RAASi therapy. There was no significant difference in the risk of recurrent hyperkalemia when RAASi was down-titrated or discontinued compared to when it was continued (aHR 1.01, 95% CI 0.97-1.05, p=0.62).

**Figure 3.**
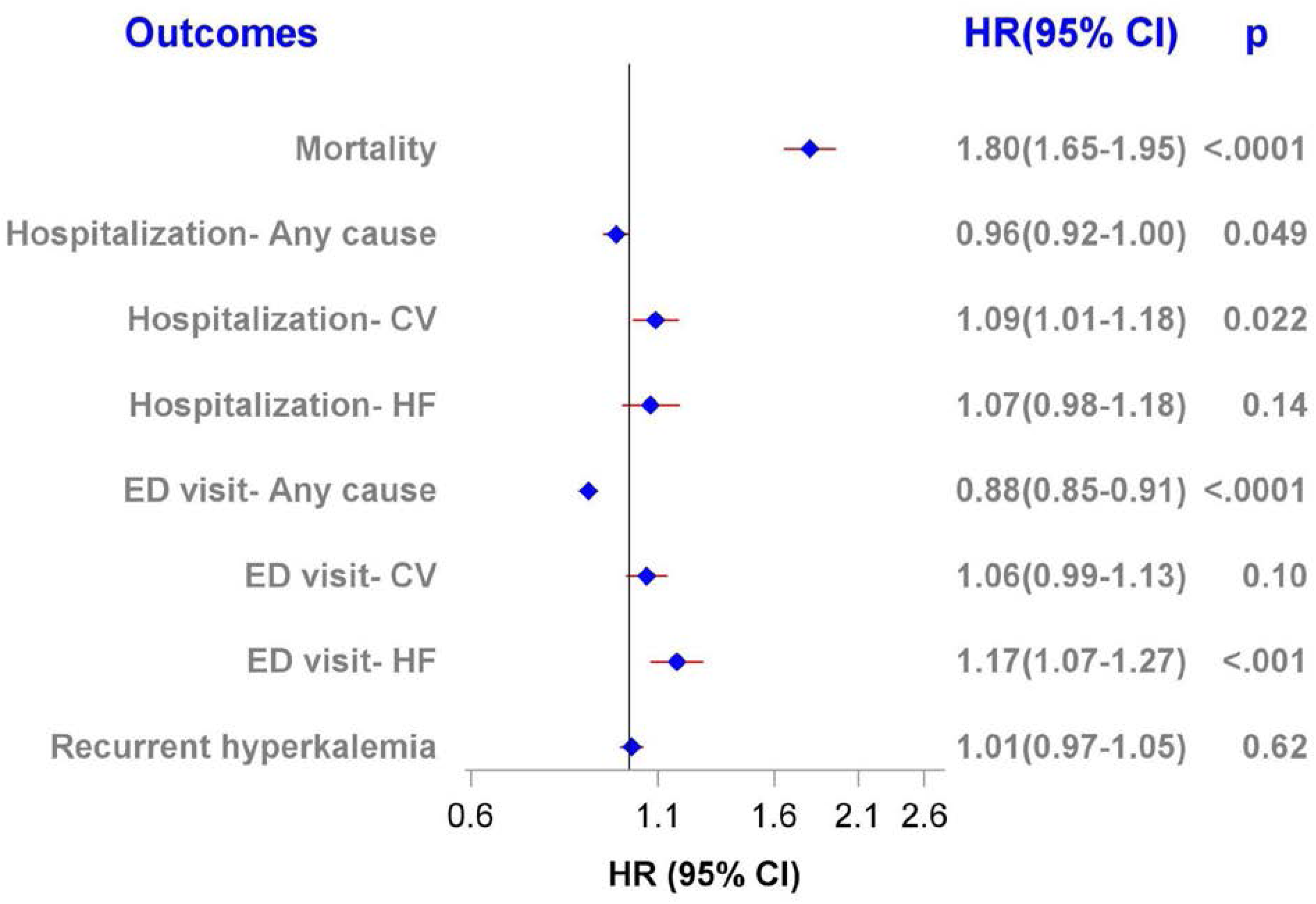
Weighted hazard ratio for discontinuation or down-titration of RAASi Adjusted HR of clinical outcomes associated with discontinuation or down-titration of RAASi over a median follow-up of 1.4 (IQR 0.7-2.0) years. RAASi, renin-angiotensin-aldosterone system inhibitors; CV, cardiovascular; ED, emergency department; HF, heart failure; HR, hazard ratio; CI, confidence interval

## Discussion

In this population-based cohort study of 7527 patients with HF who experienced hyperkalemia events related to RAASi discontinuation, there are several key findings. First, we identified that the incidence of RASSi-associated hyperkalemia was common, occurring at a rate of 17 episodes per 100 RAASi-treatment years, and that discontinuation or down-titration of RAASi was common following a hyperkalemia event. This rate is higher than prior studies, which may be attributed to the calculation of the rate, including the duration of follow-up and the number of hyperkalemia events. The present study included single events instead of multiple events per patient. Prior studies, such as the PARADIGM-HF trial (Prospective comparison of ARNI with ACEI to Determine Impact on Global Mortality and Morbidity in Heart Failure), found that the incidence of hyperkalemia in a population receiving treatment with a RAASi was approximately 7 per 100 PY.^39^ Similarly, in a large cohort study from the United States of 48,333 patients with documented HF, approximately 20 per 100 PY developed hyperkalemia in a 3-year follow-up period.^40^

Second, when a RAASi was discontinued, it was associated with an increased risk of events, including all-cause mortality, hospitalizations for cardiovascular disease and ED visits for HF. While this may not come as a surprise given the importance of RAASi (as proven efficacious in RCTs), the increased risk even in the presence of hyperkalemia is important to consider as it relates to practice and potentially reinitiating RAASi when possible. Third, there was no difference in the risk of recurrent hyperkalemia when RAASi was down-titrated or discontinued, leading to the importance of looking at all factors that may cause hyperkalemia beyond cardiovascular disease.

### Clinical Implications

Our results have important clinical implications, given the considerable cases of hyperkalemia in HF patients, and the resulting changes in RAASi therapy. Several clinical trials, such as PARADIGM-HF, RALES (Randomized Aldactone Evaluation Study), and EMPHASIS-HF (Eplerenone in Mild Patients Hospitalization and Survival Study in Heart Failure) have demonstrated the efficacy of RAAS inhibitors in patients with HF ^29,41^. Our results demonstrate that the long-term outcomes of discontinuing RAASi therapy are associated with poor outcomes in patients with HF. To our knowledge, this is the first study to examine the associations between RAASi and hyperkalemia in patients with chronic HF. A study of the associations between incident hyperkalemia during acute HF hospitalizations and subsequent RAASi management identified that incident hyperkalemia was associated with down-titration of MRAs but patients who increased or maintained their dose of RAASi had better 180-day survival ^42^. This suggests that, when possible, healthcare providers should avoid discontinuing or down-titrating RAASi in these patients, even after an acute hyperkalemia episode.

Following the treatment of an acute episode of hyperkalemia, clinicians must decide between continuing RAASi for the long-term cardiovascular benefits or discontinuing or down-titrating RAASi to avoid the short-term and intermediate-term consequences of hyperkalemia. Our results demonstrate that the risk of recurrent hyperkalemia is not significantly greater when RAASi are discontinued or down-titrated, which suggests that recurrent hyperkalemia is not always a direct result of RAASi therapy, and may be due to non-cardiovascular related causes. Despite the benefits, hyperkalemia remains a barrier to continuing to prescribe or maintain RAASi therapy ^43,44^.

### Limitations

Several limitations deserve consideration. Given that this is an observational study with a retrospective cohort design, there may be potential unmeasured confounders that could cause hyperkalemia, including over-the-counter medications. Additionally, the severity and quantity of other comorbidities could bias these patients toward a worse outcome. Similarly, some comorbidities or hemodynamic factors could limit the use or tolerance of RAASi, making them more likely to discontinue RAASi therapy. For example, blood pressure was not available in the data and thus we could not adjust for hypotension, a factor that could lead to the discontinuation or dose reduction of RAASi. However, the magnitude of the association and the overall sample size suggest that the association is unlikely due to residual confounding alone.^45^ The PIN database records when a medication is dispensed at the pharmacy; however, it does not provide information on the actual prescription from the physicians or whether the patient is taking medication. Around one-sixth of patients display primary nonadherence by not taking their prescription to be dispensed or consuming the dispensed meds. We assumed patients remained on the same dose until the end of supply or the next dispense (in 30 days from the hyperkalemia episode) since it was not feasible from the existing database to determine the temporary discontinuation/dose change. This might underestimate the dose changes or discontinuation of treatment. We only included dispenses from the first treatment date. As a result, our analysis may miss dose changes if another dispense occurred in 30 days and underestimate the number of increased or reduced doses. Finally, information regarding the reason for discontinuation of RAASi was unavailable.

## Conclusions

In summary, the current study represents an evaluation of the association between dose changes of RAASi after an episode of hyperkalemia in patients with HF and highlights the frequency of hyperkalemia in this population. Our results evaluated the effect of discontinuation or down titration of RAASi in patients with HF after an episode of hyperkalemia. Discontinuation or down titration of RAASi therapy was associated with an increased risk of all-cause mortality, hospitalizations for cardiovascular disease, and HF-related ED visits, and there was no significant difference in recurrent hyperkalemia episodes in this group. This study demonstrates the importance of monitoring hyperkalemia in HF patients; however, clinicians should be judicious in deciding to discontinue RAASi therapy.

### Clinical Perspectives

#### Clinical Competencies

The current study provides evidence of the association between discontinuation and down-titration of RAASi in patients with chronic heart failure who have experienced an episode of hyperkalemia. The study demonstrates the importance of monitoring hyperkalemia in this population, but the added importance of evaluating changes to RAASi therapy. This evidence should be considered when deciding to discontinue or dose-change RAASi, and when to reinitiate therapy in patients with heart failure, in spite of hyperkalemia.

#### Translational Outlook

While the study offers some evidence on RAASi therapy dose change and hyperkalemia, the study is limited by several factors, such as the observational nature of the study. It cannot account for all potential factors that could cause hyperkalemia, and cannot capture reasons why RAASi therapy was discontinued. Despite this, clinicians may be more cautious in dose changes of therapy, and consider more frequent monitoring and follow-up of this patient population. Current evidence supports the use of GDMT including RAASi in this population, but limited data exists on the titration of therapy in the context of adverse side effects. Future research may include RCTs that explore the effect of dose-reduction or discontinuation of RAASi compared to continuation in patients with HF who have experienced hyperkalemia. Future studies may also strengthen the evidence in the current population of patients with chronic heart failure.

## Data Availability

To request authorization to obtain these data by direct access, contact.

## Acknowledgements

Data was extracted from the Alberta Health Services Enterprise Data Warehouse with support provided by AbSPORU Data and Research Services platform, which is funded by CIHR, Alberta Innovates, University Hospital Foundation, University of Alberta, University of Calgary and Alberta Health Services. The interpretation and conclusions contained herein are those of the researchers and do not necessarily represent the views of Alberta Health Services or any of the funders.

## Abbreviations

RAASi: renin-angiotensin-aldosterone system inhibitors
GDMT: guideline-directed medical therapy
AHS: Alberta Health Services
ULI: unique lifetime identifier
NACRS: National Ambulatory Care Reporting System
DAD: Discharge Abstract Database
CCI: Canadian Classification of Health Intervention
PIN: Pharmaceutical Information Network
DIN: Drug Identification Number

## Central Illustration

Illustration of the objectives, cohort selection design and key results of the study regarding the impact of hyperkalemia on RAASI therapy and the association with clinical outcomes. CV, cardiovascular; HF, heart failure; RAASi, renin-angiotensin-aldosterone system inhibitors

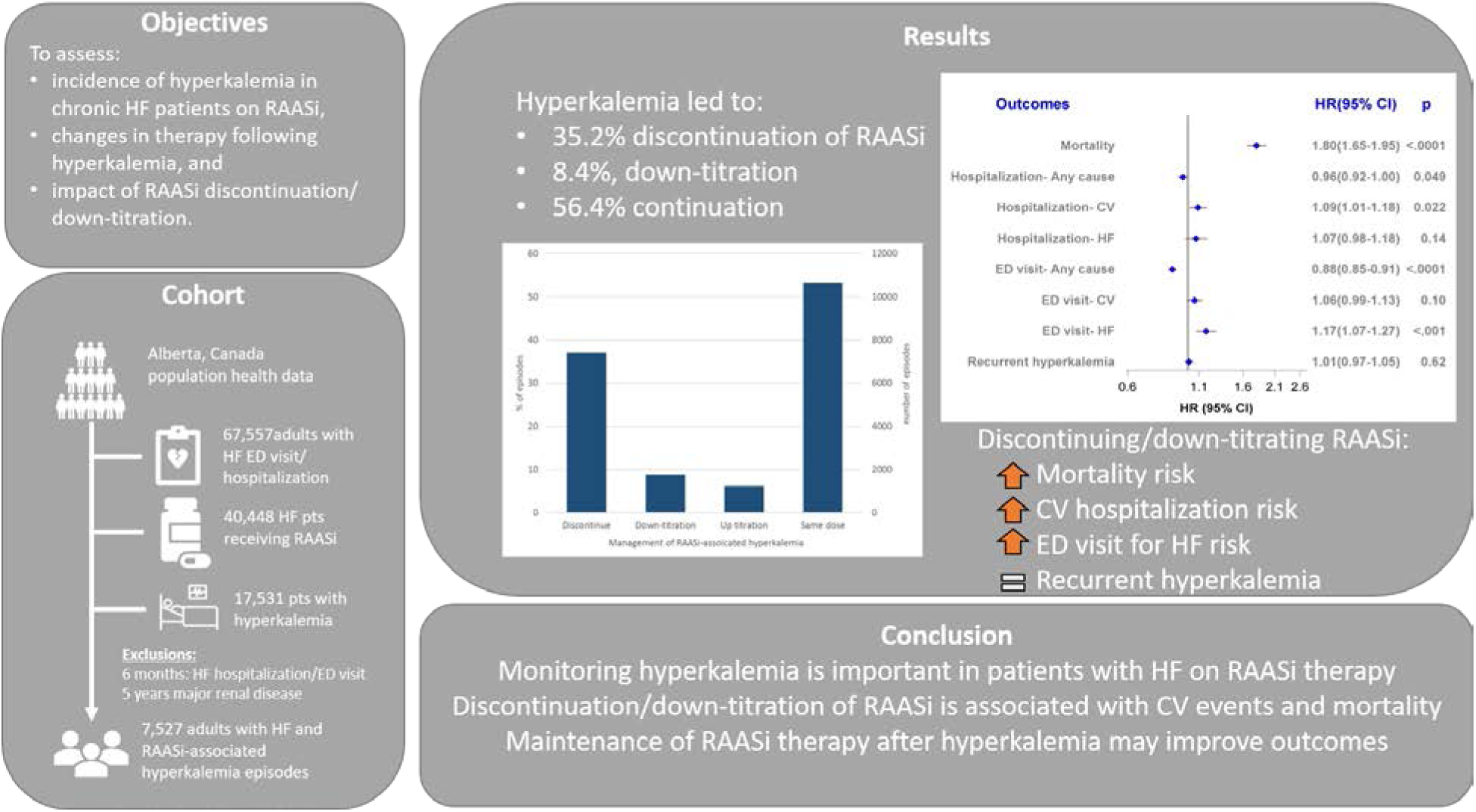

## References

[1] Tran DT, Ohinmaa A, Thanh NX, Howlett JG, Ezekowitz JA, McAlister FA, et al. The current and future financial burden of hospital admissions for heart failure in Canada: a cost analysis. CMAJ Open. 2016;4(3):E365–E370.

[2] Gislason GH, Rasmussen JN, Abildstrom SZ, Schramm TK, Hansen ML, Buch P, et al. Persistent use of evidence-based pharmacotherapy in heart failure is associated with improved outcomes. Circulation. 2007;116(7):737–744.

[3] Neubauer S, Schilling T, Zeidler J, Lange A, Engel S, Linder R, et al. [Impact of guideline adherence on mortality in treatment of left heart failure]. Herz. 2016;41(7):614–624.

[4] CONSENSUS Trial Study Group. Effects of enalapril on mortality in severe congestive heart failure. Results of the Cooperative North Scandinavian Enalapril Survival Study (CONSENSUS). N Engl J Med. 1987;316(23):1429–1435.

[5] Køber L, Torp-Pedersen C, Carlsen JE, Bagger H, Eliasen P, Lyngborg K, et al. A clinical trial of the angiotensin-converting-enzyme inhibitor trandolapril in patients with left ventricular dysfunction after myocardial infarction. Trandolapril Cardiac Evaluation (TRACE) Study Group. N Engl J Med. 1995;333(25):1670–1676.

[6] SOLVD Investigators, Yusuf S, Pitt B, Davis CE, Hood WB, Cohn JN. Effect of enalapril on survival in patients with reduced left ventricular ejection fractions and congestive heart failure. N Engl J Med. 1991;325(5):293–302.

[7] Garg R, Yusuf S. Overview of randomized trials of angiotensin-converting enzyme inhibitors on mortality and morbidity in patients with heart failure. Collaborative Group on ACE Inhibitor Trials. JAMA. 1995;273(18):1450–1456.

[8] SOLVD Investigators, Yusuf S, Pitt B, Davis CE, Hood WB Jr, Cohn JN. Effect of enalapril on mortality and the development of heart failure in asymptomatic patients with reduced left ventricular ejection fractions. N Engl J Med. 1992;327(10):685–691.

9. [9] Effect of ramipril on mortality and morbidity of survivors of acute myocardial infarction with clinical evidence of heart failure. The Acute Infarction Ramipril Efficacy (AIRE) Study Investigators. Lancet. 1993;342(8875):821–828.

[10] Pfeffer MA, McMurray JJV, Velazquez EJ, Rouleau JL, Køber L, Maggioni AP, et al. Valsartan, captopril, or both in myocardial infarction complicated by heart failure, left ventricular dysfunction, or both. N Engl J Med. 2003;349(20):1893–1906.

[11] Cohn JN, Tognoni G, Valsartan Heart Failure Trial Investigators. A randomized trial of the angiotensin-receptor blocker valsartan in chronic heart failure. N Engl J Med. 2001;345(23):1667–1675.

[12] Granger CB, McMurray JJV, Yusuf S, Held P, Michelson EL, Olofsson B, et al. Effects of candesartan in patients with chronic heart failure and reduced left-ventricular systolic function intolerant to angiotensin-converting-enzyme inhibitors: the CHARM-Alternative trial. Lancet. 2003;362(9386):772–776.

[13] Lee VC, Rhew DC, Dylan M, Badamgarav E, Braunstein GD, Weingarten SR. Meta-analysis: angiotensin-receptor blockers in chronic heart failure and high-risk acute myocardial infarction. Ann Intern Med. 2004;141(9):693–704.

[14] Rosano GMC, Tamargo J, Kjeldsen KP, Lainscak M, Agewall S, Anker SD, et al. Expert consensus document on the management of hyperkalaemia in patients with cardiovascular disease treated with renin angiotensin aldosterone system inhibitors: coordinated by the Working Group on Cardiovascular Pharmacotherapy of the European Society of Cardiology. Eur Heart J Cardiovasc Pharmacother. 2018;4(3):180–188.

[15] Aldahl M, Jensen ASC, Davidsen L, Eriksen MA, Møller Hansen S, Nielsen BJ, et al. Associations of serum potassium levels with mortality in chronic heart failure patients. Eur Heart J. 2017;38(38):2890–2896.

[16] Ponikowski P, Voors AA, Anker SD, Bueno H, Cleland JGF, Coats AJS, et al. 2016 ESC Guidelines for the diagnosis and treatment of acute and chronic heart failure: The Task Force for the diagnosis and treatment of acute and chronic heart failure of the European Society of Cardiology (ESC). Developed with the special contribution of the Heart Failure Association (HFA) of the ESC. Eur J Heart Fail. 2016;18(8):891–975.

[17] Weir MR, Rolfe M. Potassium homeostasis and renin-angiotensin-aldosterone system inhibitors. Clin J Am Soc Nephrol. 2010;5(3):531–548.

[18] Gjesing A, Schou M, Torp-Pedersen C, Køber L, Gustafsson F, Hildebrandt P, et al. Patient adherence to evidence-based pharmacotherapy in systolic heart failure and the transition of follow-up from specialized heart failure outpatient clinics to primary care. Eur J Heart Fail. 2013;15(6):671–678.

[19] Trevisan M, de Deco P, Xu H, Evans M, Lindholm B, Bellocco R, et al. Incidence, predictors and clinical management of hyperkalaemia in new users of mineralocorticoid receptor antagonists. Eur J Heart Fail. 2018;20(8):1217–1226.

[20] Savarese G, Carrero JJ, Pitt B, Anker SD, Rosano GMC, Dahlström U, et al. Factors associated with underuse of mineralocorticoid receptor antagonists in heart failure with reduced ejection fraction: an analysis of 11 215 patients from the Swedish Heart Failure Registry. Eur J Heart Fail. 2018;20(9):1326–1334.

[21] Rossignol P, Lainscak M, Crespo-Leiro MG, Laroche C, Piepoli MF, Filippatos G, et al. Unravelling the interplay between hyperkalaemia, renin-angiotensin-aldosterone inhibitor use and clinical outcomes. Data from 9222 chronic heart failure patients of the ESC-HFA- EORP Heart Failure Long-Term Registry. Eur J Heart Fail. 2020;22(8):1378–1389.

[22] Linde C, Bakhai A, Furuland H, Evans M, McEwan P, Ayoubkhani D, et al. Real-World Associations of Renin-Angiotensin-Aldosterone System Inhibitor Dose, Hyperkalemia, and Adverse Clinical Outcomes in a Cohort of Patients With New-Onset Chronic Kidney Disease or Heart Failure in the United Kingdom. J Am Heart Assoc. 2019;8(22):e012655.

[23] Schmidt M, Mansfield KE, Bhaskaran K, Nitsch D, Sørensen HT, Smeeth L, et al. Adherence to guidelines for creatinine and potassium monitoring and discontinuation following renin-angiotensin system blockade: a UK general practice-based cohort study. BMJ Open. 2017;7(1):e012818.

[24] Maggioni AP, Anker SD, Dahlström U, Filippatos G, Ponikowski P, Zannad F, et al. Are hospitalized or ambulatory patients with heart failure treated in accordance with European Society of Cardiology guidelines? Evidence from 12,440 patients of the ESC Heart Failure Long-Term Registry. Eur J Heart Fail. 2013;15(10):1173–1184.

[25] Rossignol P, Ménard J, Fay R, Gustafsson F, Pitt B, Zannad F. Eplerenone survival benefits in heart failure patients post-myocardial infarction are independent from its diuretic and potassium-sparing effects. Insights from an EPHESUS (Eplerenone Post-Acute Myocardial Infarction Heart Failure Efficacy and Survival Study) substudy. J Am Coll Cardiol. 2011;58(19):1958–1966.

[26] Vardeny O, Wu DH, Desai A, Rossignol P, Zannad F, Pitt B, et al. Influence of baseline and worsening renal function on efficacy of spironolactone in patients With severe heart failure: insights from RALES (Randomized Aldactone Evaluation Study). J Am Coll Cardiol. 2012;60(20):2082–2089.

[27] Eschalier R, McMurray JJV, Swedberg K, van Veldhuisen DJ, Krum H, Pocock SJ, et al. Safety and efficacy of eplerenone in patients at high risk for hyperkalemia and/or worsening renal function: analyses of the EMPHASIS-HF study subgroups (Eplerenone in Mild Patients Hospitalization And SurvIval Study in Heart Failure). J Am Coll Cardiol. 2013;62(17):1585–1593.

[28] Epstein M, Reaven NL, Funk SE, McGaughey KJ, Oestreicher N, Knispel J. Evaluation of the treatment gap between clinical guidelines and the utilization of renin-angiotensin-aldosterone system inhibitors. Am J Manag Care. 2015;21(11 Suppl):S212–S220.

[29] Rossignol P, Dobre D, McMurray JJV, Swedberg K, Krum H, van Veldhuisen DJ, et al. Incidence, determinants, and prognostic significance of hyperkalemia and worsening renal function in patients with heart failure receiving the mineralocorticoid receptor antagonist eplerenone or placebo in addition to optimal medical therapy: results from the Eplerenone in Mild Patients Hospitalization and Survival Study in Heart Failure (EMPHASIS-HF). Circ Heart Fail. 2014;7(1):51–58.

[30] Bakal JA, McAlister FA, Liu W, Ezekowitz JA. Heart failure re-admission: measuring the ever shortening gap between repeat heart failure hospitalizations. PLoS One. 2014;9(9):e106494.

[31] Ezekowitz JA, Kaul P, Bakal JA, Quan H, McAlister FA. Trends in heart failure care: has the incident diagnosis of heart failure shifted from the hospital to the emergency department and outpatient clinics? Eur J Heart Fail. 2011;13(2):142–147.

[32] Quan H, Parsons GA, Ghali WA. Validity of information on comorbidity derived rom ICD-9- CCM administrative data. Med Care. 2002;40(8):675–685.

[33] Hemmelgarn BR, Clement F, Manns BJ, Klarenbach S, James MT, Ravani P, et al. Overview of the Alberta Kidney Disease Network. BMC Nephrol. 2009;10:30.

[34] Sepehrvand N, Bakal JA, Lin M, McAlister F, Wesenberg JC, Ezekowitz JA. Factors Associated With Natriuretic Peptide Testing in Patients Presenting to Emergency Departments With Suspected Heart Failure. Can J Cardiol. 2016;32(8):986.e1-e8.

[35] Ghimire A, Fine N, Ezekowitz JA, Howlett J, Youngson E, McAlister FA. Frequency, predictors, and prognosis of ejection fraction improvement in heart failure: an echocardiogram-based registry study. Eur Heart J. 2019;40(26):2110–2117.

[36] Quan H, Khan N, Hemmelgarn BR, Tu K, Chen G, Campbell N, et al. Validation of a case definition to define hypertension using administrative data. Hypertension. 2009;54(6):1423–1428.

[37] Quan H, Sundararajan V, Halfon P, Fong A, Burnand B, Luthi JC, et al. Coding algorithms for defining comorbidities in ICD-9-CM and ICD-10 administrative data. Med Care. 2005;43(11):1130–1139.

[38] Imai K, Ratkovic M. Covariate Balancing Propensity Score. J R Stat Soc Series B Stat Methodol. 2013;76(1):243–263.

[39] McMurray JJV, Packer M, Desai AS, Gong J, Lefkowitz MP, Rizkala AR, et al. Angiotensin-neprilysin inhibition versus enalapril in heart failure. N Engl J Med. 2014;371(11):993–1004.

[40] Muhlestein JB, Kammerer J, Bair TL, Knowlton KU, Le VT, Anderson JL, et al. Frequency and clinical impact of hyperkalaemia within a large, modern, real-world heart failure population. ESC Heart Fail. 2021;8(1):691–696.

[41] Pitt B, Zannad F, Remme WJ, Cody R, Castaigne A, Perez A, et al. The effect of spironolactone on morbidity and mortality in patients with severe heart failure. Randomized Aldactone Evaluation Study Investigators. N Engl J Med. 1999;341(10):709–717.

[42] Hyperkalemia and Treatment With RAAS Inhibitors During Acute Heart Failure Hospitalizations and Their Association With Mortality. JACC: Heart Failure. 2019;7(11):970–979.

[43] Epstein M, Alvarez PJ, Reaven NL, Funk SE, McGaughey KJ, Brenner MS, et al. Evaluation of clinical outcomes and costs based on prescribed dose level of renin-angiotensin-aldosterone system inhibitors. Am J Manag Care. 2016;22(11 Suppl):s311–s324.

[44] Vijayakumar S, Butler J, Bakris GL. Barriers to guideline mandated renin–angiotensin inhibitor use: focus on hyperkalaemia. Eur Heart J Suppl. 2019;21(Supplement_A):A20–A27.

[45] Zeitouny S, Cheng L, Wong ST, Tadrous M, McGrail K, Law MR. Prevalence and predictors of primary nonadherence to medications prescribed in primary care. CMAJ. 2023;195(30):E1000–E1009.

