## Supplemental Materials for "Hyperkalemia-related Heart Failure Therapy Discontinuation and the Association with Outcomes in Patients with Heart Failure"

**Supplemental Figure 1**. Cohort selection

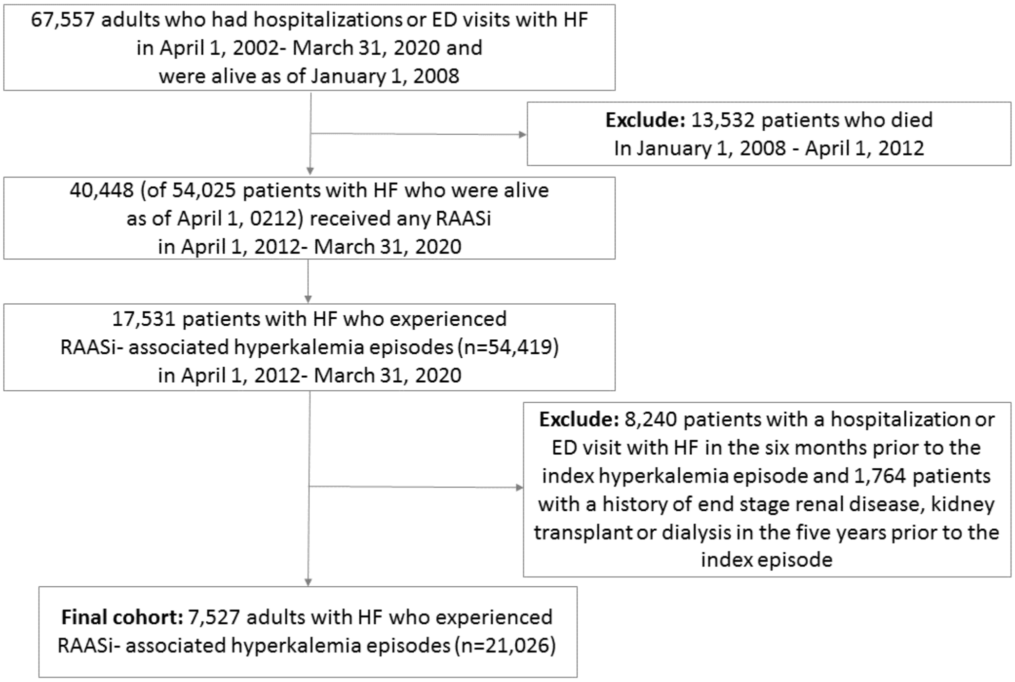


ED – Emergency department; HF – Heart failure; RAASi - Renin-angiotensin-aldosterone system inhibitors

**Supplemental Table 1**. Formulas to convert to the equivalent doses for the corresponding drug classes

| | |
|---|---|
| ***Angiotensin-converting enzyme inhibitors converted to 'lisinopril equivalents' (LisEquiv)***<br><br>LisEquiv = Lisinopril TDD<br><br>LisEquiv = Ramapril TDD *2<br><br>LisEquiv = Enalapril TDD<br><br>LisEquiv = Benazapril TDD/2<br><br>LisEquiv = Captopril TDD/7.5<br><br>LisEquiv = Quinipril TDD/2<br><br>LisEquiv = Trandolopril TDD *5<br><br>LisEquiv = Fosinopril TDD/4<br><br>LisEquiv = Perindopril TDD *5<br><br>LisEquiv = Cilazapril TDD *4 | 0 mg = 0<br><br><5.0 mg = 0.5<br><br>5.0 – 9.9 mg = 1<br><br>10 – 14.9 mg = 2<br><br>15 – 19.9 mg = 3<br><br>20 – 39.9 mg = 4<br><br>≥40 mg = 5 |
| ***Angiotensin II receptor blockers, converted to 'losartan equivalents' (LosEquiv)***<br><br>LosEquiv = Losartan TDD<br><br>LosEquiv = Valsartan TDD *0.6<br><br>LosEquiv = Candesartan TDD *6<br><br>LosEquiv = Irbesartan TDD *0.6<br><br>LosEquiv = Olmesartan TDD *4.8<br><br>LosEquiv = Telmisartan TDD *2.4 | 0 mg = 0<br><br><25 mg = 1<br><br>25 – 49.9 mg = 2<br><br>50 – 74.9 mg = 3<br><br>75 – 99.9 mg = 4<br><br>≥100 mg = 5 |
| ***Mineralocorticoid receptor antagonists, converted to 'spironolactone equivalents'***<br><br>***(SpiroEquiv)***<br><br>SpiroEquiv = Spironolactone TDD<br><br>SpiroEquiv = Eplererone TDD | 0 mg = 0<br><br><25 mg = 1<br><br>25 – 37.49 mg = 2<br><br>37.5 – 49.9 mg = 3 |

| | |
|---|---|
| | ≥50 mg = 4 |
| **Angiotensin receptor-neprilysin inhibitors**<br><br>Sacubitril/Valsartan TDD | 0mg = 0<br><br>≤ 50mg = 3<br><br>>50mg = 5 |

TDD – total daily dosage

**Supplemental Table 2**. ICD 9 and ICD 10 codes

| Comorbidities | ICD-10 Code | ICD-9 code |
|---|---|---|
| Diabetes | E10, E11, E12, E13, E14 | 250 |
| Hypertension | I10.x, I11.x-I13.x, I15.x | 401, 402, 403, 404, 405 |
| Dyslipidemia | E78.0-E78.5 | 272.4 |
| Coronary artery disease | I20, I21, I22, I23, I24, I25 | 410, 411, 412, 413, 414 |
| Atrial fibrillation | I48 | 427.3 |
| Peripheral vascular disease | I70.x, I71.x, I73.1, I73.8, I73.9, I77.1, I79.0, I79.2, K55.1. K55.8, K55.9, Z95.8, Z95.9 | 093.0, 437.3, 440, 441, 443.1, 443.2, 443.3, 443.4, 443.5, 443.6, 443.7, 443.8, 443.9, 447.1, 557.1, 557.9, V43.4 |
| Asthma | J45 | 493 |
| Chronic obstructive pulmonary disease | J41, J42, J43, J44, J47 | 491, 492, 494, 496 |
| Chronic Renal disease | I12.0, I13.1, N03.2-N03.7, N05.2-N05.7, N18.x, N19.x, N25.0, Z49.0-Z49.2, Z94.0, Z99.2 | 403, 404, 585, 586, 588, V56, V42.0, V45.1 |
| Ischemic stroke | I63.x, I64.x | 362.3, 433.01, 433.11, 433.21, 433.31, 433.41, 433.51, 433.61, 433.71, 433.81, 433.91, 434.01, 434.11, 434.21, 434.31, 434.41, 434.51, 434.61, 434.71, 434.81, 434.91, 436 |

| | | |
|---|---|---|
| Transient ischemic attack | G45 | 435 |
| Hemorrhagic stroke | I61 | 430, 431, 432 |
| Peripheral artery disease | I65.x, I70x-I73.x | 433, 440, 441, 442, 443 |
| Thromboembolism | I26, I801, I802, I803, I808, I809, I828, I829, I821, I822, I823, I801, I802, I803, I822, I823, I828, I829, I74 | 415, 451.1, 451.8, 451.9, 453.1, 453.2, 453.4, 453.5, 453.8, 453.9, 415, 451.1, 451.2, 453.3, 453.8, 453.9, 444 |
| Cancer | C0, C1, C2, C3, C5, C6, C7, C8, C9, C40, C41, C42, C43, C45, C46, C48, C49 | 14, 15, 16, 170, 171, 172, 174, 175, 176, 177, 178, 179, 180-189, 190-199, 200-208, 238.6, |
| Anemia | D50, D539, D649 | 280, 281.9, 285.9 |
| Depression | F204, F313, F314, F315, F32, F33, F341, F412, F432 | 296.2, 296.3, 296.5, 300.4, 309, 311 |
| Liver disease | B18, K73, K74, K700, K701, K702, K703, K709, K717, K713, K714, K715, K760, K762, K763, K764, K768, K769, Z944, K704, K711, K721, K729, K765, K766, K767, I850, I859, I864, I982, K704, K711, K721, K729, K765, K766, K767, I850, I859, I864, I982 | 070.22, 070.23, 070.32, 070.33, 070.44, 070.54, 070.6, 070.9, 570, 571, 573.3, 573.4, 573.8, 573.9, V42.7, 456.0, 456.1, 456.2, 572.2, 572.3, 572.4, 572.8, 456.0, 456.1, 456.2, 572.2, 572.3, 572.4, 572.8 |
| Dementia | F00, F01, F02, F03, F051, G30, G311 | 290, 294.1, 331.2 |
| Smoking | Z720 | 305.1 |

| Medications | ATC code |
|---|---|
| ACEi | C09A, C09B |
| ARB | C09C, C09D |
| ARNi | C09DX04 |
| MRA | C03DA |

ICD – International Classification of Disease; ACEi – angiotensin-converting enzyme inhibitors; ARB – angiotensin receptor blockers; ARNI – angiotensin receptor-neprilysin inhibitors; MRA – mineralocorticoid receptor antagonists; ATC – Anatomical Therapeutic Chemical Classification
